# Lyme disease incidence in the United States, 2016-2023

**DOI:** 10.64898/2026.08.06.26359815

**Authors:** Sheryl A. Kluberg, Sarah J. Willis, June O’Neill, Katherine Shapiro, Kevin Coughlin, Diane Emerton, Edward Rosen, Robert Jin, John Aucott, Kimberly Daniels, Shelly-Ann M. Love, Djeneba Audrey Djibo, Mano Selvan, Andrea DeVries, Qianli Ma, L. Hannah Gould, James H. Stark, Jennifer C. Moïsi, Noelle M. Cocoros

**Affiliations:** Harvard Pilgrim Health Care Institute, Boston, MA, USA; Harvard Medical School, Boston, MA, USA; Pfizer Inc., Cambridge, Massachusetts, USA; Johns Hopkins School of Medicine, Baltimore, Maryland, USA; Carelon Research, Wilmington, Delaware, USA; CVS Health, Blue Bell, Pennsylvania, USA; Humana Healthcare Research, Inc., Louisville, Kentucky, USA; Pfizer, Inc. New York, NY, USA; Pfizer Inc., Paris, France

**Author notes:** Corresponding author: Sheryl A. Kluberg, 401 Park Drive, Suite 401E, Boston, MA 02215, USA.

**Keywords:** Lyme disease, administrative claims, public health surveillance, vector borne diseases

## Abstract

Traditional surveillance underestimates Lyme disease (LD) incidence in the United States (US). We aimed to estimate national LD incidence using validated algorithms to identify LD cases in administrative claims data. We identified potential LD cases in commercial and Medicare claims, classified cases by disease stage, adjusted case counts using algorithm-specific positive predictive values, and standardized the adjusted counts to the US population.

The study population included >66 million individuals per year. After adjustment and standardization, we estimated 191.7, 26.8, and 16.6 new cases per 100,000 population in high-incidence, neighboring, and low-incidence states, respectively, with 24% of cases diagnosed with disseminated disease. The relative burden of disseminated LD was highest in low-incidence states (28.6%) and increased with age.

This study corroborates published estimates of national LD incidence and elucidates patterns of disease stage at diagnosis. The substantial burden of disseminated LD underscores the need for earlier detection and treatment of LD.

## Background

Lyme disease (LD), a bacterial infection transmitted by the bite of infected *Ixodes* ticks, is the most common vector-borne disease in the United States (US) (*1*). LD often presents as localized disease in the days or weeks after infection, marked by fever, chills, headache, muscle and joint pain, and an erythema migrans rash, though frequency of these symptoms varies substantially across patients. If left untreated, LD may progress to early disseminated disease with neurologic complications (e.g., numbness, tingling, facial palsy) or cardiac manifestations (atrioventricular block). Further progression may lead to arthritis of large joints or additional neurologic symptoms (e.g., peripheral neuropathy, encephalopathy) (*1*). In addition to its physical impact on patients, LD has substantial economic burden associated with medical visits, antibiotic treatments, and time off from work or school. Two US studies have estimated that medical costs incurred by a case of disseminated LD are at least double those associated with localized LD (*2,3*).

Diagnosing LD at any stage can be challenging, in part due to the nonspecific nature and broad range of symptoms with which a patient may present, as well as the relative complexity of diagnostic testing (*4*). Given the challenges with LD diagnosis and variations in reporting of infections across the country, the true morbidity burden of LD in the US cannot be estimated from surveillance data alone (*5–7*). To better ascertain the burden of LD in the US, the Centers for Disease Control and Prevention (CDC) evaluated the incidence of LD diagnoses in claims data, resulting in an estimated ∼476,000 cases of LD annually in 2010-2018 (*8*). However, that study was conducted prior to the availability of LD algorithm performance metrics, without data on people >65 years of age, and did not characterize the frequency of localized versus disseminated LD.

In the current study, we applied validated algorithms to administrative claims data to estimate the incidence of LD in the US from 2016 to 2023 according to region, age, and sex, with a particular focus on localized and disseminated LD incidence.

## Methods

### Study design and data sources

Five sources of health care administrative claims data were included in this study. Four national research entities affiliated with commercial insurers participated: Carelon Research, Inc., CVS Health, Humana Healthcare Research, Inc., and Optum’s de-identified Clinformatics^®^ Data Mart Database. Data from the Centers for Medicare & Medicaid Services (CMS) Medicare, fee-for-service (Parts A and B) and Medicare Advantage (Part C), were also included.

To be eligible for inclusion in the study, we required individuals to have continuous enrollment in medical and pharmacy benefits for at least one complete calendar year from 2016 through 2023, allowing 45-day gaps in coverage for commercial insurers and 30-day gaps for Medicare (due to differences in the data structure). An individual could be included in some years and not others. We also required complete data for sex, age, and state of residency, and only included individuals residing in the 50 US states. Among the participating commercial health plans, we excluded members enrolled in Medicare Advantage to avoid overlap with the CMS Medicare data.

For each year of the study period, we identified all potential incident LD cases based on the algorithms described below as well as the denominator of all eligible members who met our inclusion and exclusion criteria.

### Algorithms to identify Lyme disease cases

We applied three algorithms to identify potential LD cases (Figure 1; see Supplement Table 1 for further detail). A validated “primary” algorithm (*8–11*) and an unvalidated but widely used inpatient algorithm (*9,12,13*) identified outpatient and inpatient LD diagnoses, respectively. A validated “secondary” algorithm identified additional cases of disseminated LD (musculoskeletal, nervous system, and cardiovascular manifestations) without an LD-specific diagnosis code (*11*). While the inpatient algorithm has not been validated, it is expected to have a higher positive predictive value (PPV) than the outpatient LD diagnosis algorithm, as inpatient diagnoses are typically more reliable than outpatient diagnoses (*14–16*).

**Figure 1:**
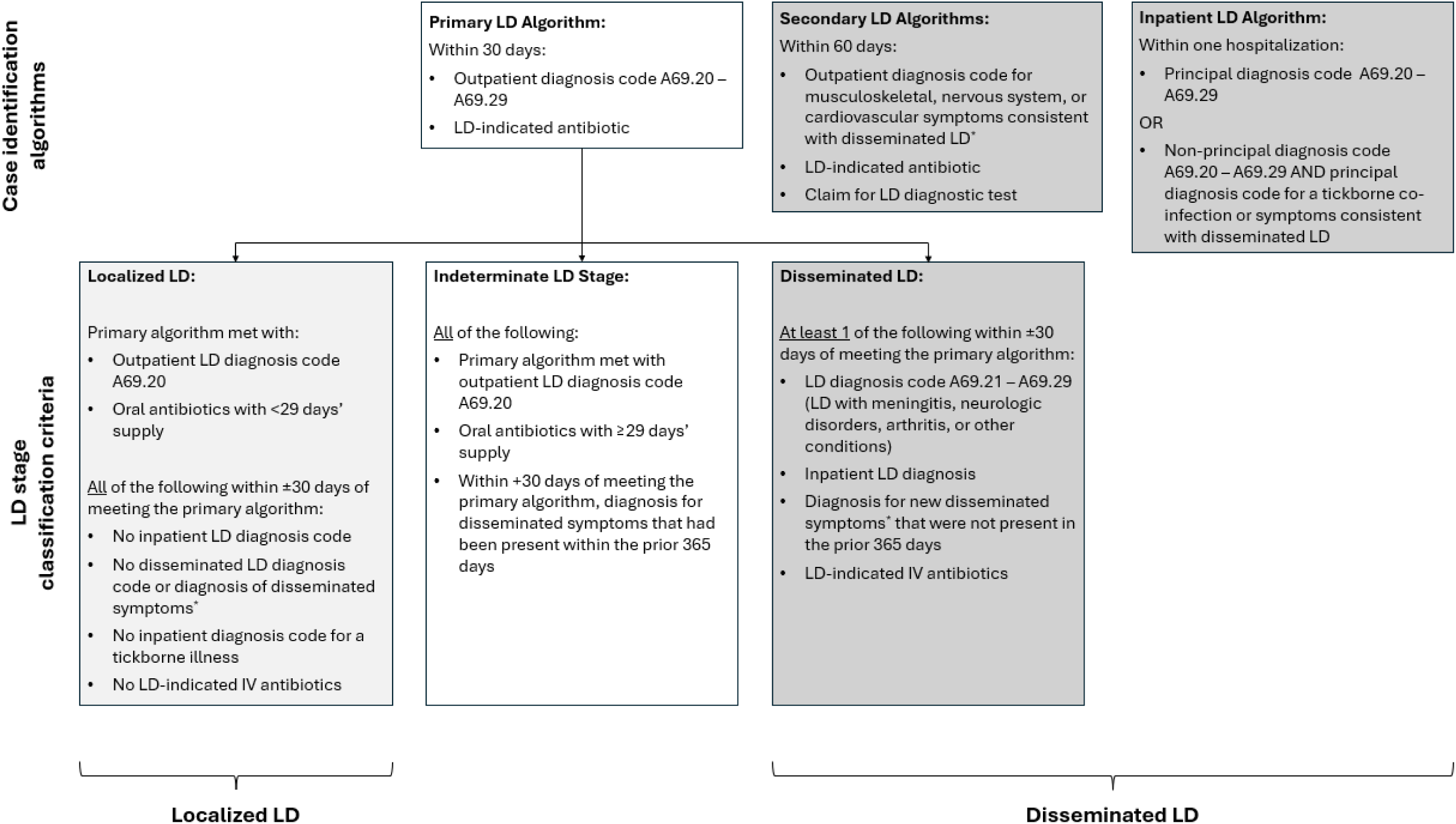
In a study estimating US Lyme disease incidence using administrative claims data, these criteria were applied to identify and describe potential Lyme disease cases. The case identification algorithms identified potential cases of Lyme disease, and the LD stage classification criteria were used to categorize cases according to the stage of disease at which the case was diagnosed. IV = intravenous; LD = Lyme disease *The following conditions defined disseminated LD symptoms: arthritis and joint effusion, arthropathy and pain in joint, encephalitis, facial palsy and facial weakness, meningitis, mononeuropathy, other cranial nerve injuries and disorders, peripheral neuropathy and polyneuropathy, atrioventricular block, carditis and myocarditis, and conduction disorders.

### LD incidence definition

To identify incident LD, we allowed up to one LD event per individual in each calendar year. If an individual met multiple algorithms in any given calendar year, priority was assigned as follows: 1) inpatient algorithm, 2) primary algorithm, 3) secondary algorithm. The first instance of the highest priority LD algorithm was selected as the LD case date. The diagnosis was considered incident if in the 365 days prior no algorithm for LD was met and fewer than three LD-specific diagnosis codes were recorded.

### Classification of Lyme disease cases by stage

After identifying potential LD cases, we classified them according to disease stage as localized, disseminated, and indeterminate. Cases meeting the primary algorithm were staged as localized or disseminated LD based on which LD diagnosis code was used, route and duration of antibiotic treatment, whether the case was hospitalized, and whether new symptoms consistent with disseminated LD were diagnosed (*17,18*). Cases identified by the primary algorithm that did not meet the localized or disseminated definition were considered indeterminate. Cases meeting the secondary and inpatient algorithms were classified as disseminated.

### Analyses

We described the demographics, applicable diagnosis codes, and outpatient pharmacy dispensings for LD treatment in the ±60 days around the case date for cases meeting each algorithm.

To estimate national LD incidence per calendar year we identified the counts of LD cases that met each of the primary, secondary, and inpatient algorithms by state, year-month, 5-year age category, and sex, summing counts across all data sources. We then adjusted observed case counts downward according to previously estimated region-specific PPVs. Algorithm validation had not been performed for low-incidence states or for the inpatient algorithm; we therefore adjusted case counts from low-incidence states by PPVs from neighboring states and applied 100% PPV for the inpatient algorithm (Table 1). Geographic regions were defined by LD burden according to the CDC’s 2023 classification (Figure 2) (*19*). According to US census data, over half of the US population (53%) resided in low-incidence states, 26% resided in high-incidence states, and 21% resided in states neighboring high-incidence states (*20*). For each stratum (state, age, sex, year), we divided the PPV-adjusted case counts by the denominator of all eligible members to calculate the stratum-specific LD incidence proportion. Finally, we calculated adjusted case counts for each stratum by applying the incidence proportion to the stratum-specific US census population size (*20,21*). To assess standardized case counts within any demographic of interest (e.g., females), by year, we summed the adjusted case counts for all rows representing that demographic (e.g., females of all ages, states) in each year. Standardized incidence per 100,000 population was calculated by dividing this total case count by the total US population for the demographic of interest in each year and multiplying by 100,000.

**Figure 2:**
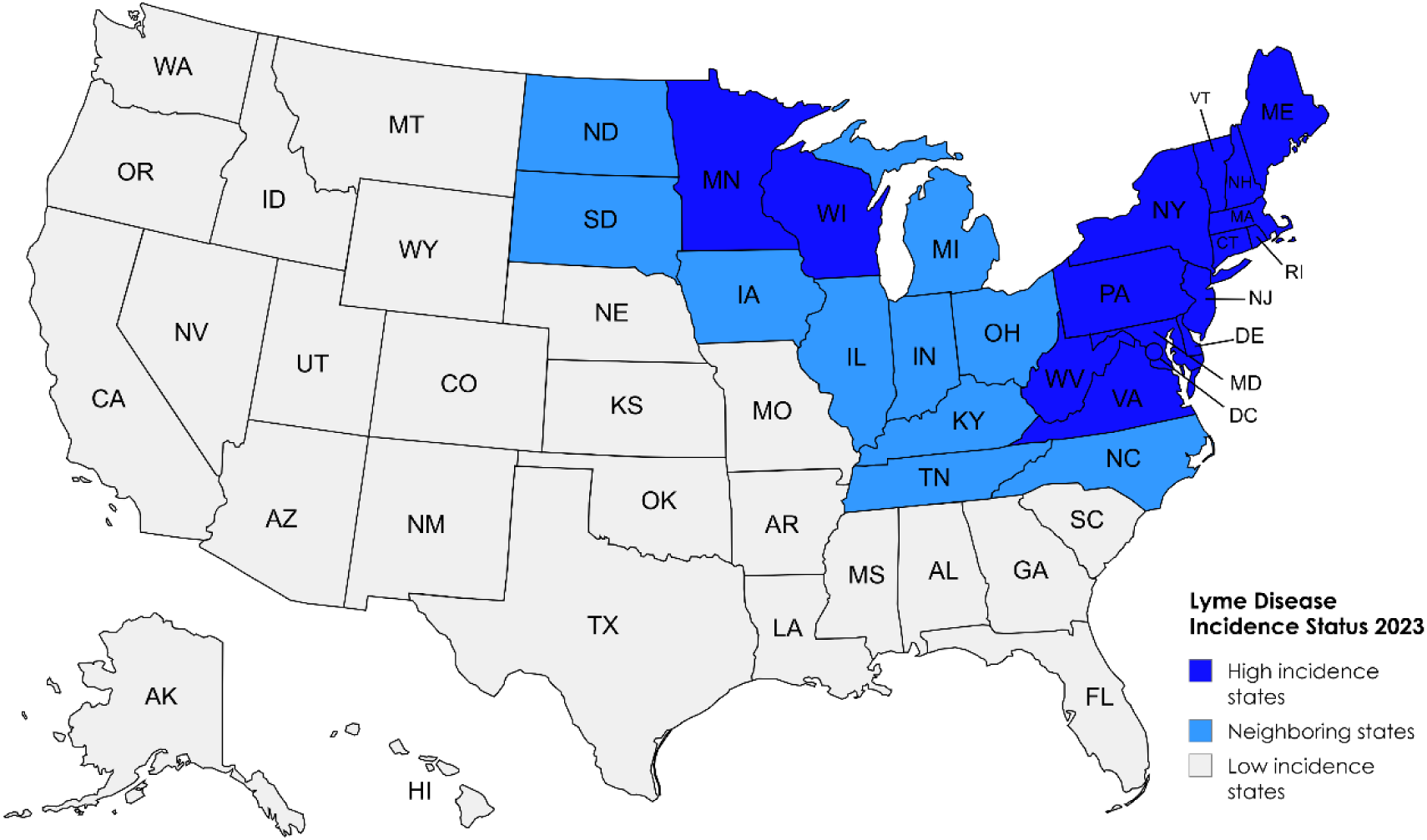
In a study estimating US Lyme disease incidence using administrative claims data, states were categorized into regions defined by level of Lyme disease activity according to the US Centers for Disease Control and Prevention (CDC). **High-incidence states** are defined by the CDC as states with an average Lyme disease incidence of ≥10 confirmed cases per 100,000 population for a period of three consecutive years.(26) **Neighboring states** are adjacent to high-incidence states and do not meet the high -incidence definition. All remaining states are classified as **low-incidence states**.

**Table 1.**
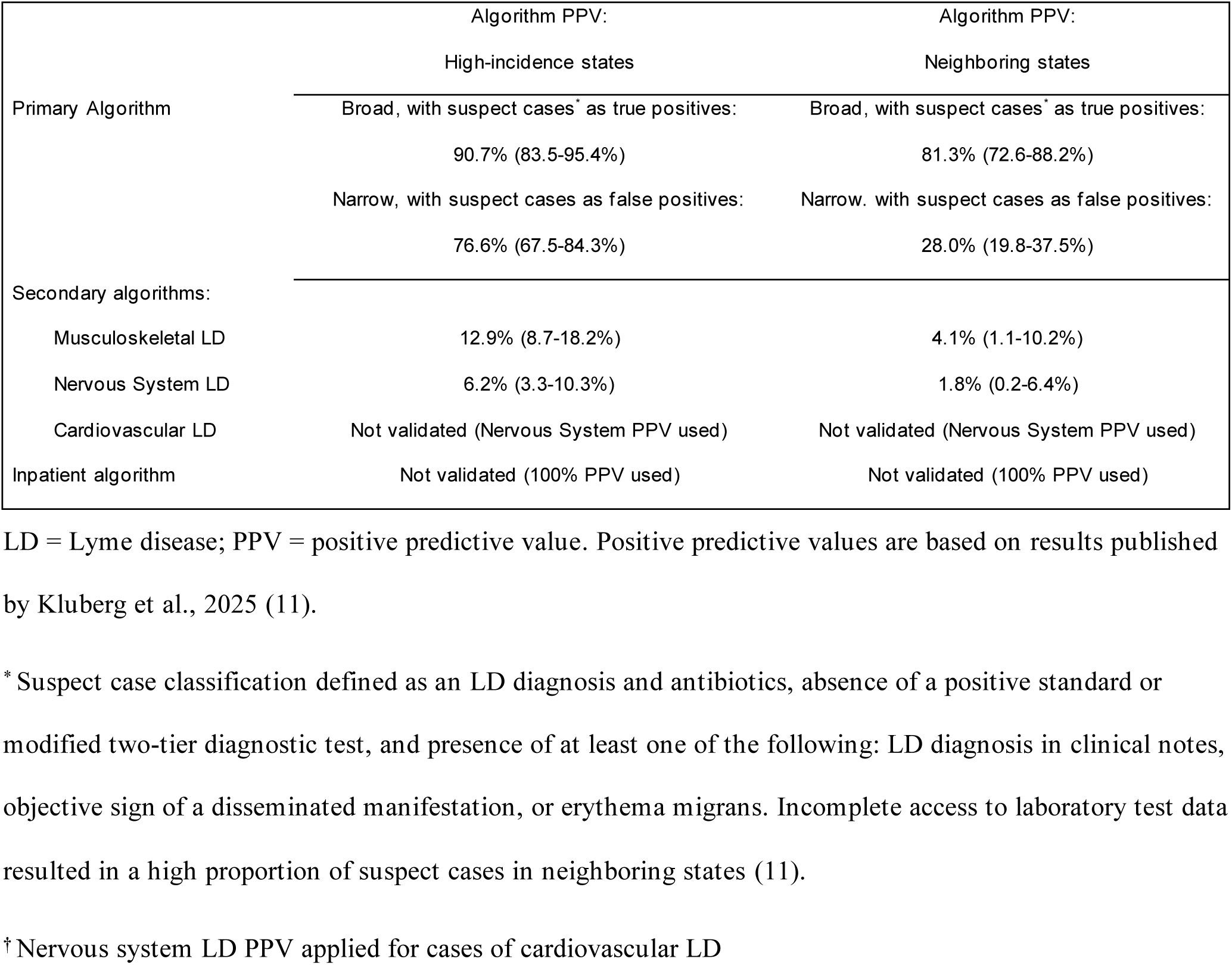
Published positive predictive values for claims-based algorithms to identify cases of Lyme disease.

|  | Algorithm PPV:<br>High-incidence states | Algorithm PPV:<br>Neighboring states |
| --- | --- | --- |
| Primary Algorithm | Broad, with suspect cases<sup>*</sup> as true positives:<br>90.7% (83.5-95.4%)<br>Narrow, with suspect cases as false positives:<br>76.6% (67.5-84.3%) | Broad, with suspect cases<sup>*</sup> as true positives:<br>81.3% (72.6-88.2%)<br>Narrow, with suspect cases as false positives:<br>28.0% (19.8-37.5%) |
| Secondary algorithms: |  |  |
| Musculoskeletal LD | 12.9% (8.7-18.2%) | 4.1% (1.1-10.2%) |
| Nervous System LD | 6.2% (3.3-10.3%) | 1.8% (0.2-6.4%) |
| Cardiovascular LD | Not validated (Nervous System PPV used) | Not validated (Nervous System PPV used) |
| Inpatient algorithm | Not validated (100% PPV used) | Not validated (100% PPV used) |

LD = Lyme disease; PPV = positive predictive value. Positive predictive values are based on results published by Kluberg et al., 2025 (11).
\* Suspect case classification defined as an LD diagnosis and antibiotics, absence of a positive standard or modified two-tier diagnostic test, and presence of at least one of the following: LD diagnosis in clinical notes, objective sign of a disseminated manifestation, or erythema migrans. Incomplete access to laboratory test data resulted in a high proportion of suspect cases in neighboring states (11).
† Nervous system LD PPV applied for cases of cardiovascular LD

We calculated an overall incidence estimate for all LD, as well as estimates by algorithm and for cases classified as localized or disseminated LD. We used the lower and upper bounds of the 95% confidence interval (CI) of the PPV to calculate bounds for standardized estimates.

Sensitivity analyses adjusted for imperfect sensitivity of the primary algorithm. We assigned the primary algorithm sensitivity as 61.5%, which translates to a multiplier of 1.626 (=1/0.615), based on a weighted average of sensitivity estimates from two recent studies using electronic health record data (*22,23*). We also applied a lower PPV for the primary algorithm in low-incidence states, recognizing that PPV is a function of disease prevalence. With no empirical estimate of algorithm PPV in low-incidence states, we applied values of 50% and 20%. We did not include cases identified by the secondary algorithms in these sensitivity analyses, as implementation of the sensitivity multiplier serves the same function as the secondary algorithms, which is to account for LD cases that did not receive a LD diagnosis code.

All analyses were conducted using SAS version 9.4 (SAS Institute Inc). The Institutional Review Board of the Harvard Pilgrim Health Care Institute deemed that this activity was not human subjects research.

## Results

### Descriptive results

Descriptive results are reported in Table 2. All five data sources combined included over 66 million members each year (except in 2023, for which 48 million members were included, as Medicare Advantage [Part C] data were not available).

**Table 2.**
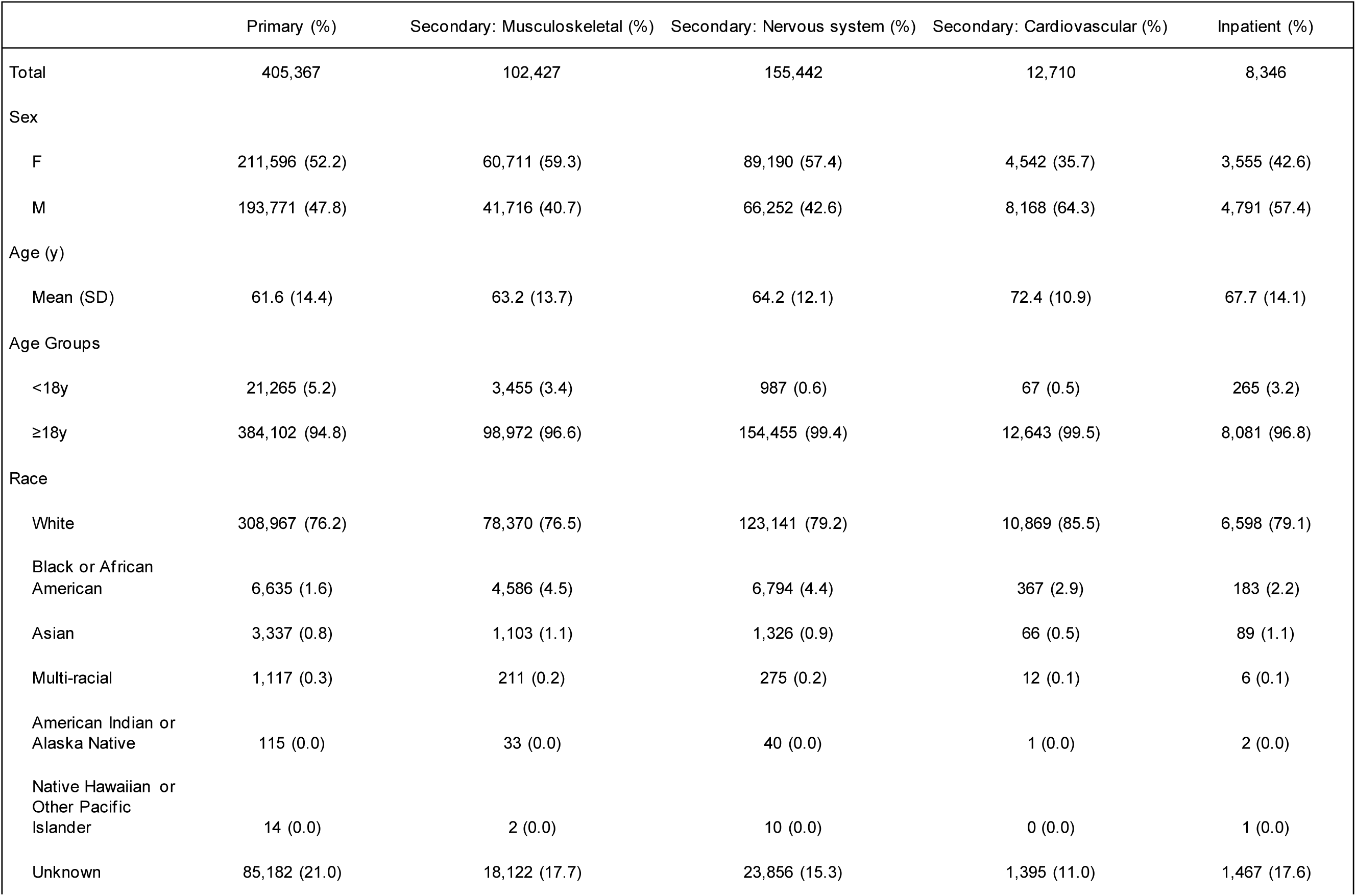
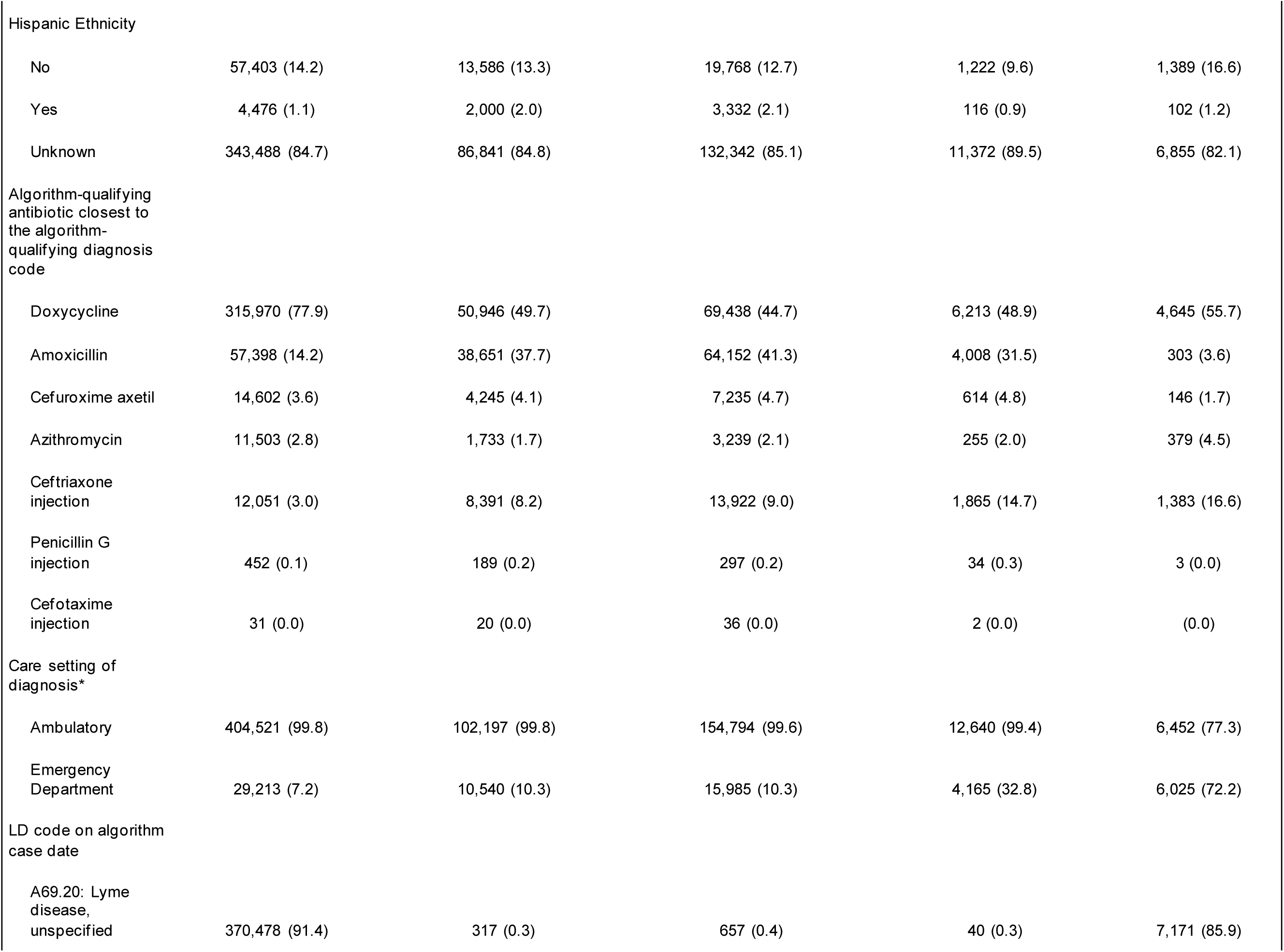
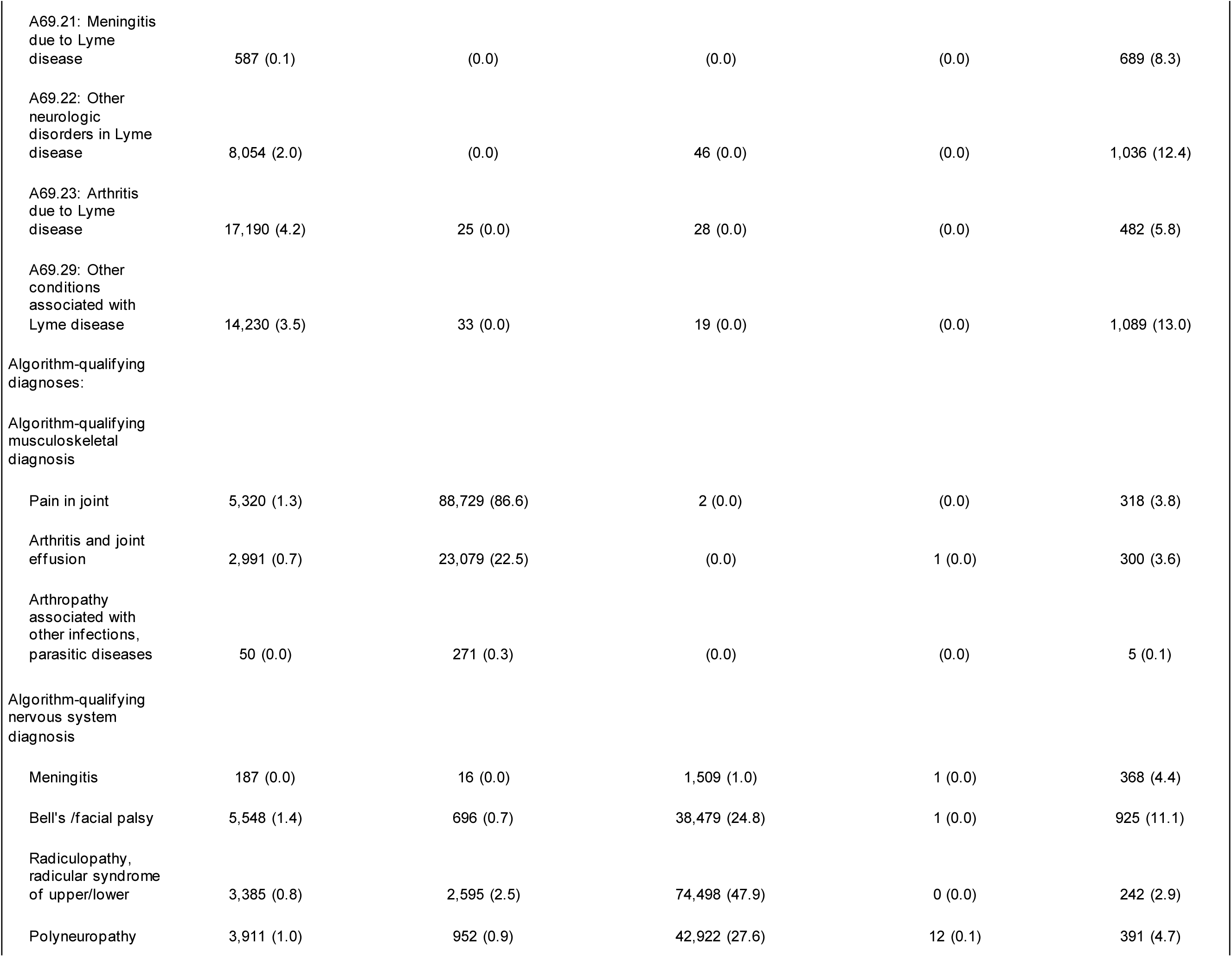
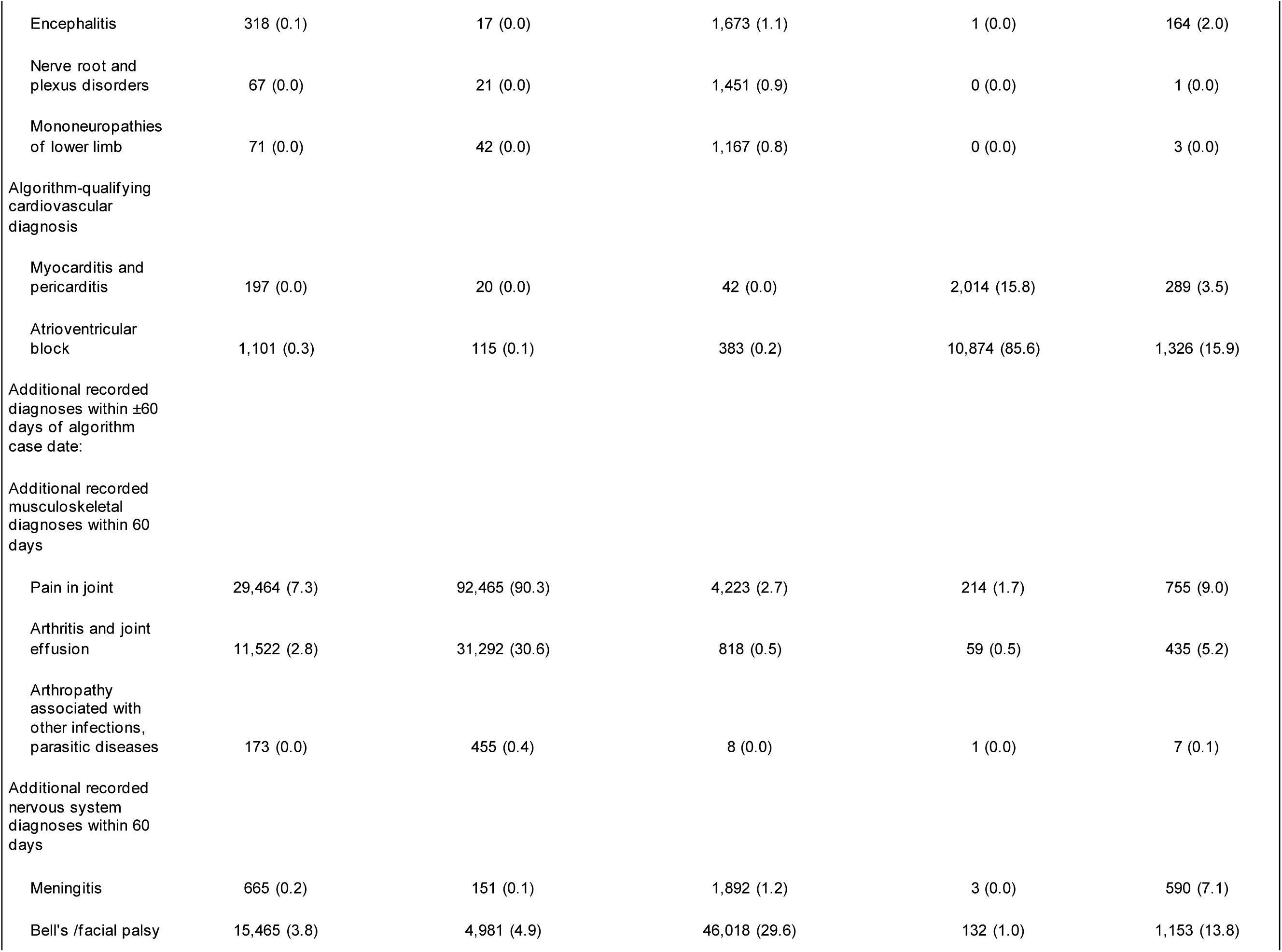
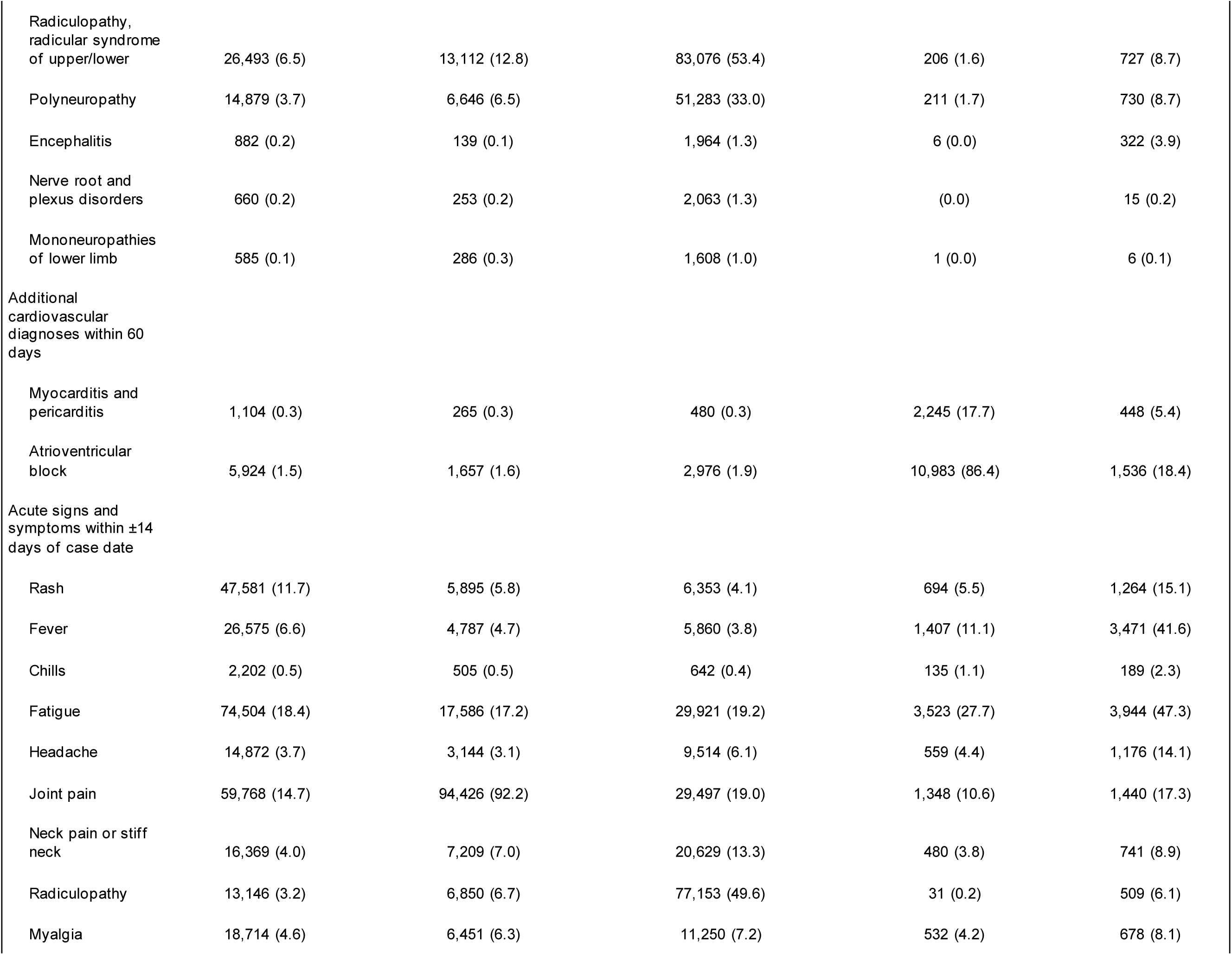
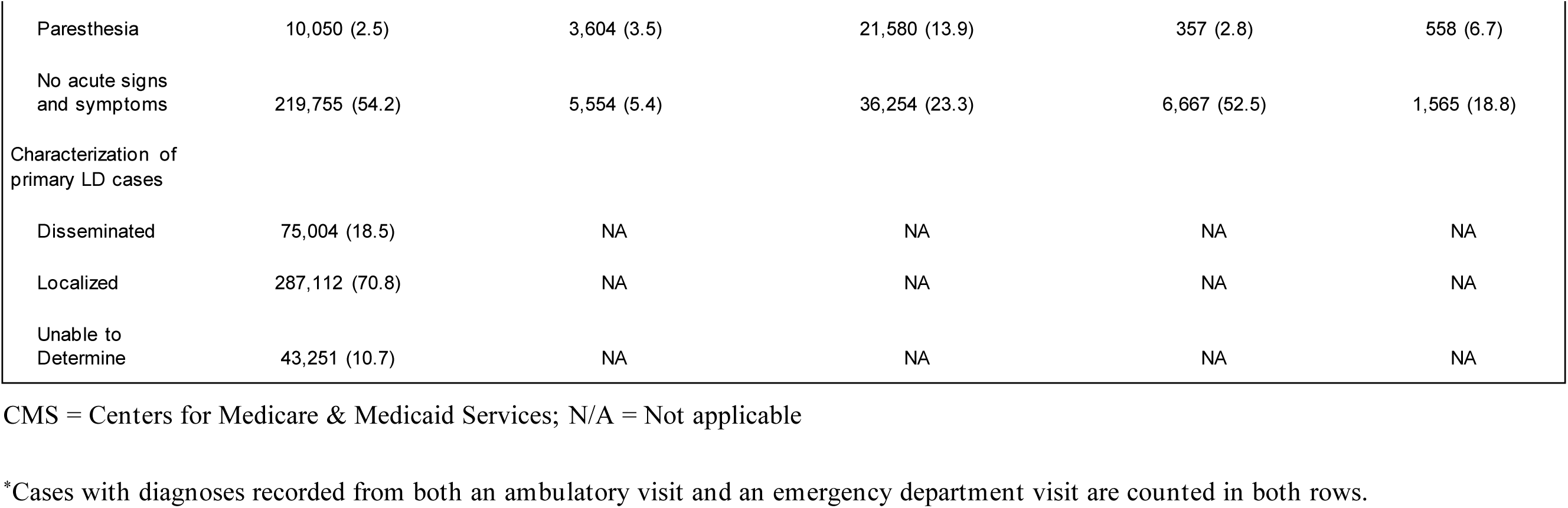
Descriptive characteristics of cases meeting the Primary, Secondary (Musculoskeletal, Nervous System, Cardiovascular), and Inpatient Lyme disease algorithms from 2016 through 2023 from four national commercial insurers and CMS Medicare

|  | Primary (%) | Secondary: Musculoskeletal (%) | Secondary: Nervous system (%) | Secondary: Cardiovascular (%) | Inpatient (%) |
| --- | --- | --- | --- | --- | --- |
| Total | 405,367 | 102,427 | 155,442 | 12,710 | 8,346 |
| Sex |  |  |  |  |  |
| F | 211,596 (52.2) | 60,711 (59.3) | 89,190 (57.4) | 4,542 (35.7) | 3,555 (42.6) |
| M | 193,771 (47.8) | 41,716 (40.7) | 66,252 (42.6) | 8,168 (64.3) | 4,791 (57.4) |
| Age (y) |  |  |  |  |  |
| Mean (SD) | 61.6 (14.4) | 63.2 (13.7) | 64.2 (12.1) | 72.4 (10.9) | 67.7 (14.1) |
| Age Groups |  |  |  |  |  |
| <18y | 21,265 (5.2) | 3,455 (3.4) | 987 (0.6) | 67 (0.5) | 265 (3.2) |
| ≥18y | 384,102 (94.8) | 98,972 (96.6) | 154,455 (99.4) | 12,643 (99.5) | 8,081 (96.8) |
| Race |  |  |  |  |  |
| White | 308,967 (76.2) | 78,370 (76.5) | 123,141 (79.2) | 10,869 (85.5) | 6,598 (79.1) |
| Black or African American | 6,635 (1.6) | 4,586 (4.5) | 6,794 (4.4) | 367 (2.9) | 183 (2.2) |
| Asian | 3,337 (0.8) | 1,103 (1.1) | 1,326 (0.9) | 66 (0.5) | 89 (1.1) |
| Multi-racial | 1,117 (0.3) | 211 (0.2) | 275 (0.2) | 12 (0.1) | 6 (0.1) |
| American Indian or Alaska Native | 115 (0.0) | 33 (0.0) | 40 (0.0) | 1 (0.0) | 2 (0.0) |
| Native Hawaiian or Other Pacific Islander | 14 (0.0) | 2 (0.0) | 10 (0.0) | 0 (0.0) | 1 (0.0) |
| Unknown | 85,182 (21.0) | 18,122 (17.7) | 23,856 (15.3) | 1,395 (11.0) | 1,467 (17.6) |
| Hispanic Ethnicity |  |  |  |  |  |
| No | 57,403 (14.2) | 13,586 (13.3) | 19,768 (12.7) | 1,222 (9.6) | 1,389 (16.6) |
| Yes | 4,476 (1.1) | 2,000 (2.0) | 3,332 (2.1) | 116 (0.9) | 102 (1.2) |
| Unknown | 343,488 (84.7) | 86,841 (84.8) | 132,342 (85.1) | 11,372 (89.5) | 6,855 (82.1) |
| Algorithm-qualifying antibiotic closest to the algorithm-qualifying diagnosis code |  |  |  |  |  |
| Doxycycline | 315,970 (77.9) | 50,946 (49.7) | 69,438 (44.7) | 6,213 (48.9) | 4,645 (55.7) |
| Amoxicillin | 57,398 (14.2) | 38,651 (37.7) | 64,152 (41.3) | 4,008 (31.5) | 303 (3.6) |
| Cefuroxime axetil | 14,602 (3.6) | 4,245 (4.1) | 7,235 (4.7) | 614 (4.8) | 146 (1.7) |
| Azithromycin | 11,503 (2.8) | 1,733 (1.7) | 3,239 (2.1) | 255 (2.0) | 379 (4.5) |
| Ceftriaxone injection | 12,051 (3.0) | 8,391 (8.2) | 13,922 (9.0) | 1,865 (14.7) | 1,383 (16.6) |
| Penicillin G injection | 452 (0.1) | 189 (0.2) | 297 (0.2) | 34 (0.3) | 3 (0.0) |
| Cefotaxime injection | 31 (0.0) | 20 (0.0) | 36 (0.0) | 2 (0.0) | (0.0) |
| Care setting of diagnosis* |  |  |  |  |  |
| Ambulatory | 404,521 (99.8) | 102,197 (99.8) | 154,794 (99.6) | 12,640 (99.4) | 6,452 (77.3) |
| Emergency Department | 29,213 (7.2) | 10,540 (10.3) | 15,985 (10.3) | 4,165 (32.8) | 6,025 (72.2) |
| LD code on algorithm case date |  |  |  |  |  |
| A69.20: Lyme disease, unspecified | 370,478 (91.4) | 317 (0.3) | 657 (0.4) | 40 (0.3) | 7,171 (85.9) |
| A69.21: Meningitis due to Lyme disease | 587 (0.1) | (0.0) | (0.0) | (0.0) | 689 (8.3) |
| A69.22: Other neurologic disorders in Lyme disease | 8,054 (2.0) | (0.0) | 46 (0.0) | (0.0) | 1,036 (12.4) |
| A69.23: Arthritis due to Lyme disease | 17,190 (4.2) | 25 (0.0) | 28 (0.0) | (0.0) | 482 (5.8) |
| A69.29: Other conditions associated with Lyme disease | 14,230 (3.5) | 33 (0.0) | 19 (0.0) | (0.0) | 1,089 (13.0) |
| Algorithm-qualifying diagnoses: |  |  |  |  |  |
| Algorithm-qualifying musculoskeletal diagnosis |  |  |  |  |  |
| Pain in joint | 5,320 (1.3) | 88,729 (86.6) | 2 (0.0) | (0.0) | 318 (3.8) |
| Arthritis and joint effusion | 2,991 (0.7) | 23,079 (22.5) | (0.0) | 1 (0.0) | 300 (3.6) |
| Arthropathy associated with other infections, parasitic diseases | 50 (0.0) | 271 (0.3) | (0.0) | (0.0) | 5 (0.1) |
| Algorithm-qualifying nervous system diagnosis |  |  |  |  |  |
| Meningitis | 187 (0.0) | 16 (0.0) | 1,509 (1.0) | 1 (0.0) | 368 (4.4) |
| Bell's /facial palsy | 5,548 (1.4) | 696 (0.7) | 38,479 (24.8) | 1 (0.0) | 925 (11.1) |
| Radiculopathy, radicular syndrome of upper/lower | 3,385 (0.8) | 2,595 (2.5) | 74,498 (47.9) | 0 (0.0) | 242 (2.9) |
| Polyneuropathy | 3,911 (1.0) | 952 (0.9) | 42,922 (27.6) | 12 (0.1) | 391 (4.7) |
| Encephalitis | 318 (0.1) | 17 (0.0) | 1,673 (1.1) | 1 (0.0) | 164 (2.0) |
| Nerve root and plexus disorders | 67 (0.0) | 21 (0.0) | 1,451 (0.9) | 0 (0.0) | 1 (0.0) |
| Mononeuropathies of lower limb | 71 (0.0) | 42 (0.0) | 1,167 (0.8) | 0 (0.0) | 3 (0.0) |
| Algorithm-qualifying cardiovascular diagnosis |  |  |  |  |  |
| Myocarditis and pericarditis | 197 (0.0) | 20 (0.0) | 42 (0.0) | 2,014 (15.8) | 289 (3.5) |
| Atrioventricular block | 1,101 (0.3) | 115 (0.1) | 383 (0.2) | 10,874 (85.6) | 1,326 (15.9) |
| Additional recorded diagnoses within ±60 days of algorithm case date: |  |  |  |  |  |
| Additional recorded musculoskeletal diagnoses within 60 days |  |  |  |  |  |
| Pain in joint | 29,464 (7.3) | 92,465 (90.3) | 4,223 (2.7) | 214 (1.7) | 755 (9.0) |
| Arthritis and joint effusion | 11,522 (2.8) | 31,292 (30.6) | 818 (0.5) | 59 (0.5) | 435 (5.2) |
| Arthropathy associated with other infections, parasitic diseases | 173 (0.0) | 455 (0.4) | 8 (0.0) | 1 (0.0) | 7 (0.1) |
| Additional recorded nervous system diagnoses within 60 days |  |  |  |  |  |
| Meningitis | 665 (0.2) | 151 (0.1) | 1,892 (1.2) | 3 (0.0) | 590 (7.1) |
| Bell's /facial palsy | 15,465 (3.8) | 4,981 (4.9) | 46,018 (29.6) | 132 (1.0) | 1,153 (13.8) |
| Radiculopathy, radicular syndrome of upper/lower | 26,493 (6.5) | 13,112 (12.8) | 83,076 (53.4) | 206 (1.6) | 727 (8.7) |
| Polyneuropathy | 14,879 (3.7) | 6,646 (6.5) | 51,283 (33.0) | 211 (1.7) | 730 (8.7) |
| Encephalitis | 882 (0.2) | 139 (0.1) | 1,964 (1.3) | 6 (0.0) | 322 (3.9) |
| Nerve root and plexus disorders | 660 (0.2) | 253 (0.2) | 2,063 (1.3) | (0.0) | 15 (0.2) |
| Mononeuropathies of lower limb | 585 (0.1) | 286 (0.3) | 1,608 (1.0) | 1 (0.0) | 6 (0.1) |
| Additional cardiovascular diagnoses within 60 days |  |  |  |  |  |
| Myocarditis and pericarditis | 1,104 (0.3) | 265 (0.3) | 480 (0.3) | 2,245 (17.7) | 448 (5.4) |
| Atrioventricular block | 5,924 (1.5) | 1,657 (1.6) | 2,976 (1.9) | 10,983 (86.4) | 1,536 (18.4) |
| Acute signs and symptoms within ±14 days of case date |  |  |  |  |  |
| Rash | 47,581 (11.7) | 5,895 (5.8) | 6,353 (4.1) | 694 (5.5) | 1,264 (15.1) |
| Fever | 26,575 (6.6) | 4,787 (4.7) | 5,860 (3.8) | 1,407 (11.1) | 3,471 (41.6) |
| Chills | 2,202 (0.5) | 505 (0.5) | 642 (0.4) | 135 (1.1) | 189 (2.3) |
| Fatigue | 74,504 (18.4) | 17,586 (17.2) | 29,921 (19.2) | 3,523 (27.7) | 3,944 (47.3) |
| Headache | 14,872 (3.7) | 3,144 (3.1) | 9,514 (6.1) | 559 (4.4) | 1,176 (14.1) |
| Joint pain | 59,768 (14.7) | 94,426 (92.2) | 29,497 (19.0) | 1,348 (10.6) | 1,440 (17.3) |
| Neck pain or stiff neck | 16,369 (4.0) | 7,209 (7.0) | 20,629 (13.3) | 480 (3.8) | 741 (8.9) |
| Radiculopathy | 13,146 (3.2) | 6,850 (6.7) | 77,153 (49.6) | 31 (0.2) | 509 (6.1) |
| Myalgia | 18,714 (4.6) | 6,451 (6.3) | 11,250 (7.2) | 532 (4.2) | 678 (8.1) |
| Paresthesia | 10,050 (2.5) | 3,604 (3.5) | 21,580 (13.9) | 357 (2.8) | 558 (6.7) |
| No acute signs and symptoms | 219,755 (54.2) | 5,554 (5.4) | 36,254 (23.3) | 6,667 (52.5) | 1,565 (18.8) |
| Characterization of primary LD cases |  |  |  |  |  |
| Disseminated | 75,004 (18.5) | NA | NA | NA | NA |
| Localized | 287,112 (70.8) | NA | NA | NA | NA |
| Unable to Determine | 43,251 (10.7) | NA | NA | NA | NA |

CMS = Centers for Medicare & Medicaid Services; N/A = Not applicable
\*Cases with diagnoses recorded from both an ambulatory visit and an emergency department visit are counted in both rows.

From 2016 through 2023, an average of 85,537 LD cases per year were identified across all algorithms prior to any PPV adjustment or standardization. A total of 405,367 cases, or an average of 50,671 cases per year, were identified via the primary algorithm with an LD code. Nearly all (91.4%) received the diagnosis code A69.20, “Lyme Disease, unspecified.” Among cases meeting the primary algorithm, 287,112 (70.8%) were classified as localized LD and 75,004 (18.5%) as disseminated LD; the remainder were indeterminate.

Secondary algorithms for disseminated disease without an LD-specific diagnosis code identified 102,427 total cases (12,803 annual average) with musculoskeletal symptoms; the most common algorithm-qualifying diagnosis among these cases was joint pain (86.6%). The algorithm identified 155,442 cases with nervous system symptoms via diagnosis codes (19,430 annual average) based on radiculopathy (47.9%), polyneuropathy (27.6%), and facial palsy (24.8%) diagnoses. A total of 12,710 cases met the secondary algorithm for cardiovascular LD (1,589 annual average), among whom 85.6% qualified based on a diagnosis of atrioventricular block.

The inpatient algorithm identified a total of 8,346 LD cases (1,043 annual average). The most common coded symptoms were fatigue (47.3%), fever (41.6%), joint pain (17.3%), rash (15.1%), and headache (14.1%). Supplement Table 2 describes characteristics of cases classified as disseminated versus localized disease stage among those that were identified via the primary algorithm.

### Lyme disease incidence

As noted above, we estimated an annual average of 85,537 LD cases across all algorithms; adjustment by PPVs resulted in an annual average of 48,126 LD cases using the “broad” PPV for the primary algorithm (with suspect cases considered true positives). After standardizing these case counts to the US population by state, age, and sex, we estimated a national average of 212,628 cases of new-onset LD each year (range: 180,556 – 251,101), corresponding to an incidence of 64.6 cases per 100,000 population (Table 3). Annually, on average, 140,886 (66.3%) cases were classified as localized LD and 49,937 (23.5%) as disseminated LD. The remainder (21,805 (10.3%)) did not fit either category. Applying the “narrow” PPV for the primary algorithm (with suspect cases considered false positives), we estimated an annual average of 159,004 total cases (range: 134,039 – 188,365; incidence: 48.3 cases per 100,000 population). Standardized national LD case counts trended slightly downward from 2016 through 2023, although this linear trend was not statistically significant (average annual decrease by 5,209 cases; p = 0.12) (Figure 3). The remainder of this section describes results calculated based on the broad PPV.

**Figure 3:**
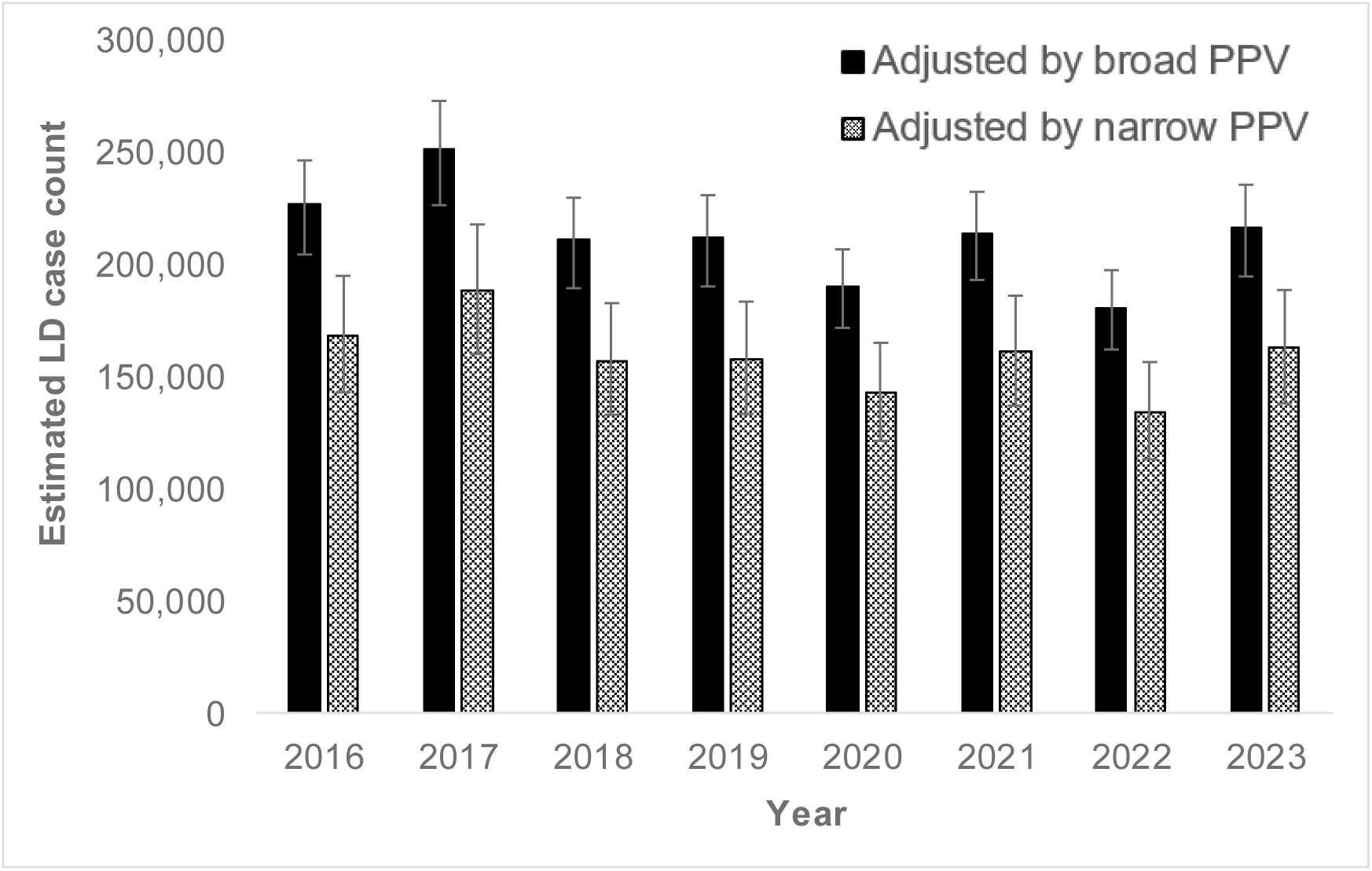
This figure depicts the annual burden of Lyme disease (LD) in the United States estimated from administrative claims data. Estimated LD case counts are adjusted for the positive predictive values (PPVs) of case-identification algorithms and standardized to the US population, with error bars representing the LD burden based on the lower and upper bounds of the 95% confidence interval of the PPV.

**Table 3.**
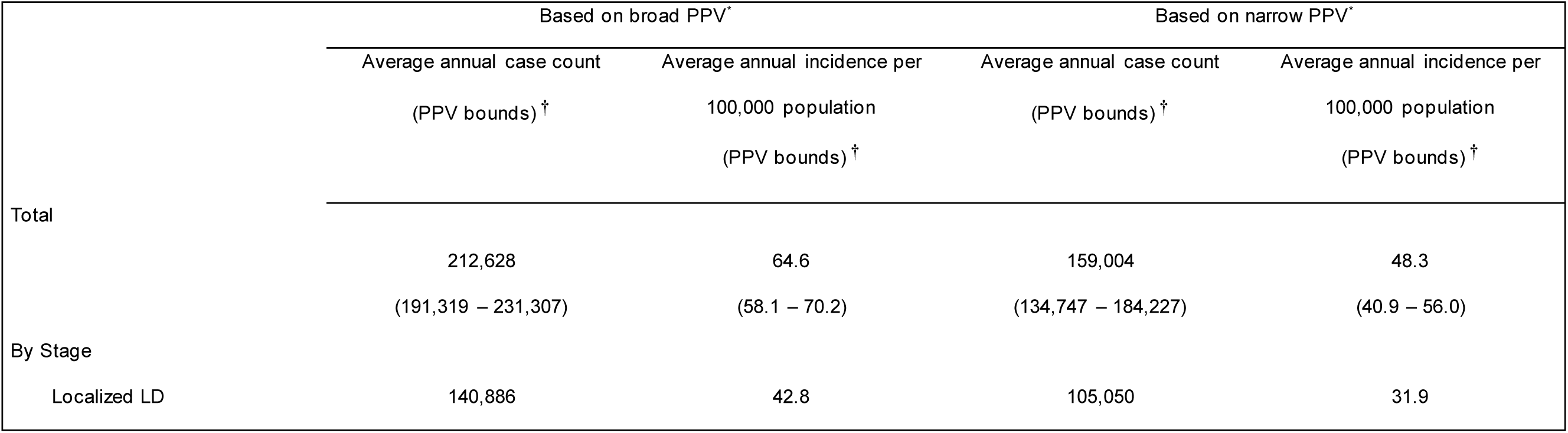
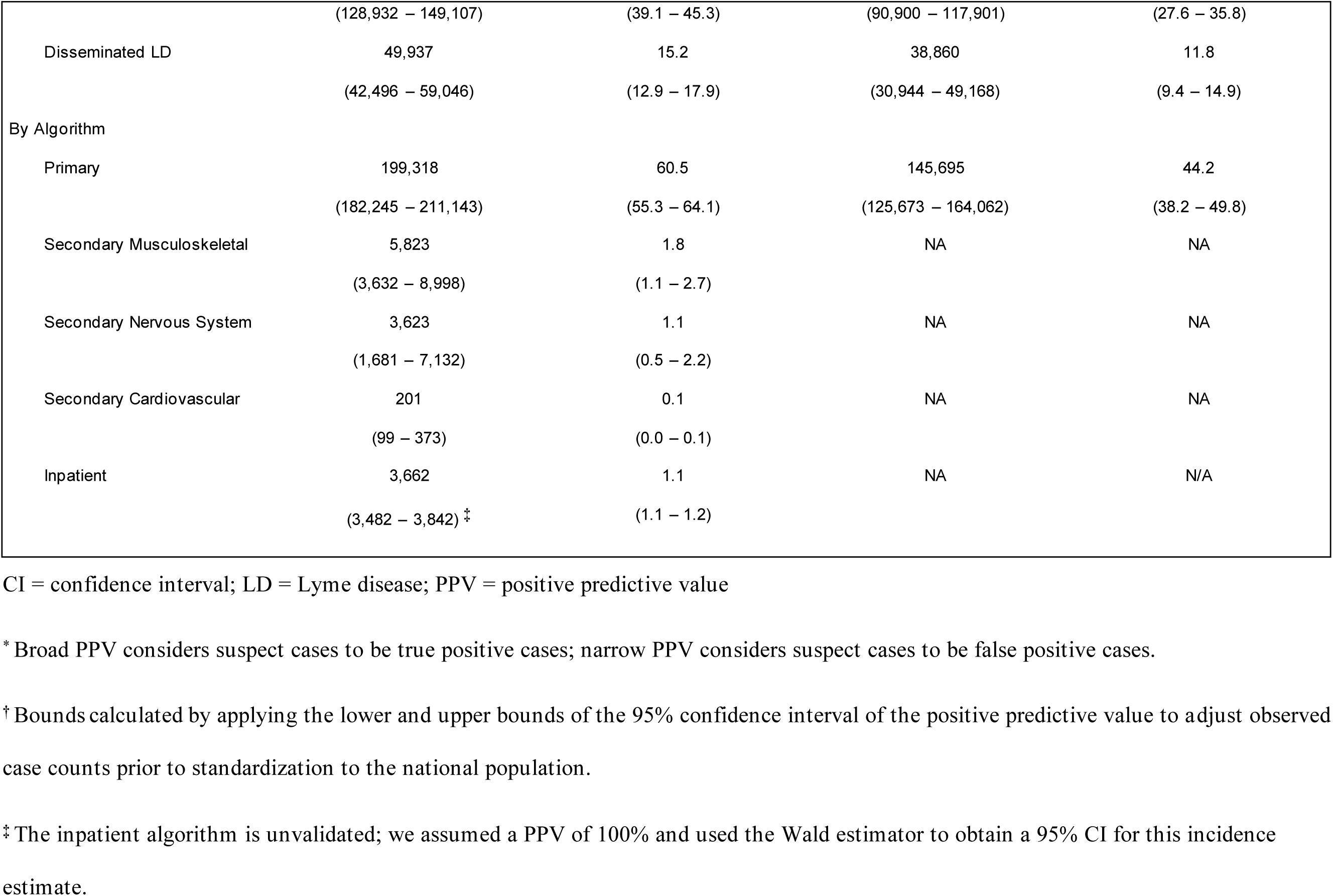
Estimated average annual case count and incidence of Lyme disease per 100,000 population overall, by stage, and by algorithm, 2016 – 2023.

|  | Based on broad PPV* |  | Based on narrow PPV* |  |
| --- | --- | --- | --- | --- |
|  | Average annual case count | Average annual incidence per | Average annual case count | Average annual incidence per |
|  | (PPV bounds) † | 100,000 population | (PPV bounds) † | 100,000 population |
|  |  | (PPV bounds) † |  | (PPV bounds) † |
| Total | 212,628<br>(191,319 – 231,307) | 64.6<br>(58.1 – 70.2) | 159,004<br>(134,747 – 184,227) | 48.3<br>(40.9 – 56.0) |
| By Stage |  |  |  |  |
| Localized LD | 140,886 | 42.8 | 105,050 | 31.9 |
|  | (128,932 – 149,107) | (39.1 – 45.3) | (90,900 – 117,901) | (27.6 – 35.8) |
| Disseminated LD | 49,937 | 15.2 | 38,860 | 11.8 |
|  | (42,496 – 59,046) | (12.9 – 17.9) | (30,944 – 49,168) | (9.4 – 14.9) |
| By Algorithm |  |  |  |  |
| Primary | 199,318 | 60.5 | 145,695 | 44.2 |
|  | (182,245 – 211,143) | (55.3 – 64.1) | (125,673 – 164,062) | (38.2 – 49.8) |
| Secondary Musculoskeletal | 5,823 | 1.8 | NA | NA |
|  | (3,632 – 8,998) | (1.1 – 2.7) |  |  |
| Secondary Nervous System | 3,623 | 1.1 | NA | NA |
|  | (1,681 – 7,132) | (0.5 – 2.2) |  |  |
| Secondary Cardiovascular | 201 | 0.1 | NA | NA |
|  | (99 – 373) | (0.0 – 0.1) |  |  |
| Inpatient | 3,662 | 1.1 | NA | N/A |
|  | (3,482 – 3,842)<sup>‡</sup> | (1.1 – 1.2) |  |  |

CI = confidence interval; LD = Lyme disease; PPV = positive predictive value
\* Broad PPV considers suspect cases to be true positive cases; narrow PPV considers suspect cases to be false positive cases.
† Bounds calculated by applying the lower and upper bounds of the 95% confidence interval of the positive predictive value to adjust observed case counts prior to standardization to the national population.
‡ The inpatient algorithm is unvalidated; we assumed a PPV of 100% and used the Wald estimator to obtain a 95% CI for this incidence estimate.

Of the 212,628 estimated LD cases per year, 165,312 (77.7%) occurred in high -incidence states, 18,220 (8.6%) in neighboring states, and 29,096 (13.7%) in low-incidence states (Table 4). A sensitivity analysis adjusting for 61.5% sensitivity of the primary algorithm, using the neighboring state primary algorithm PPV (81.3%) for low-incidence states, estimated 327,794 national LD cases per year. Varying the primary algorithm PPV for low-incidence states to 50% and 20% resulted in estimates of 310,295 and 293,516 total LD cases per year, respectively (Appendix Table 1).

**Table 4.**
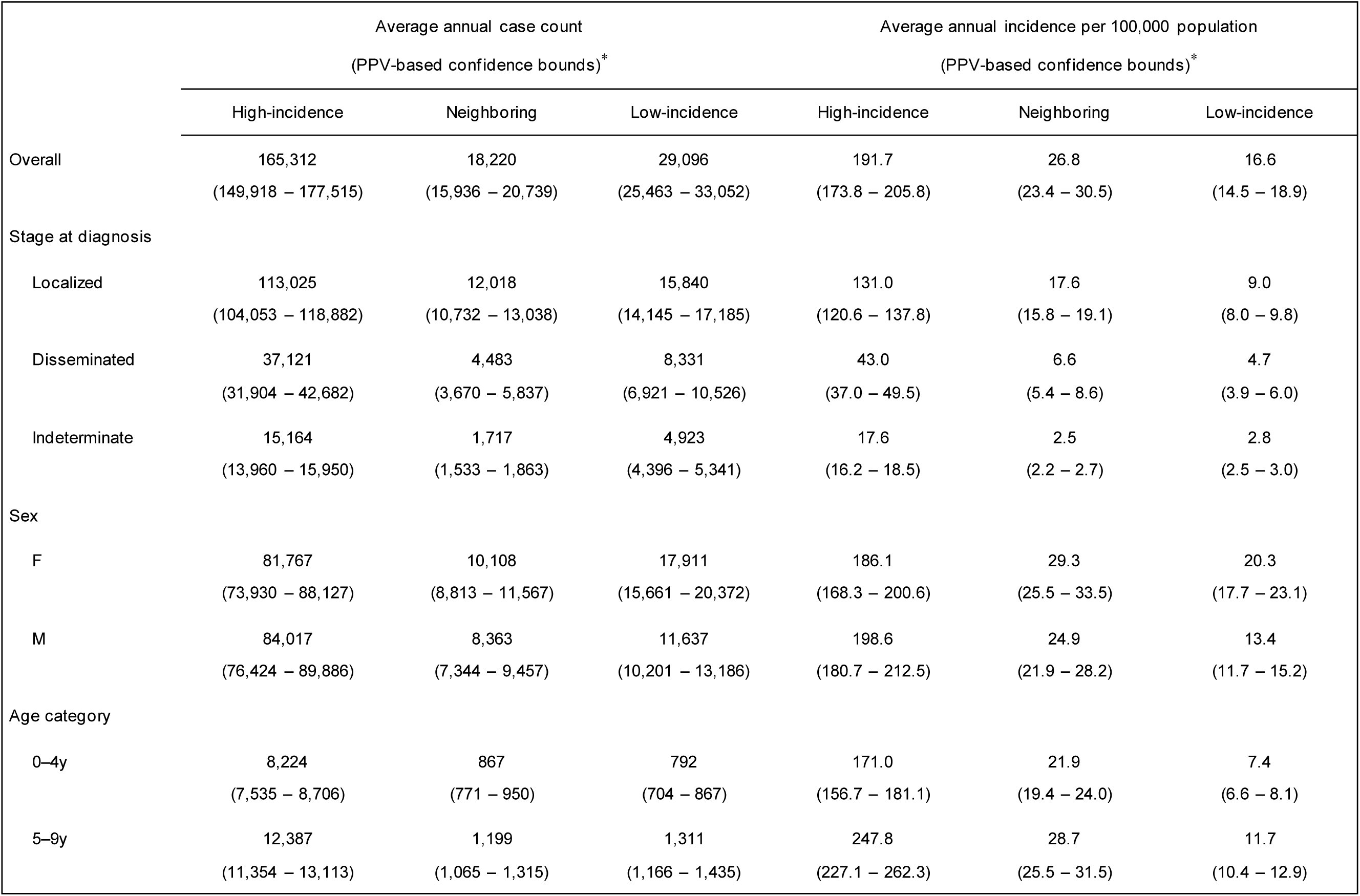
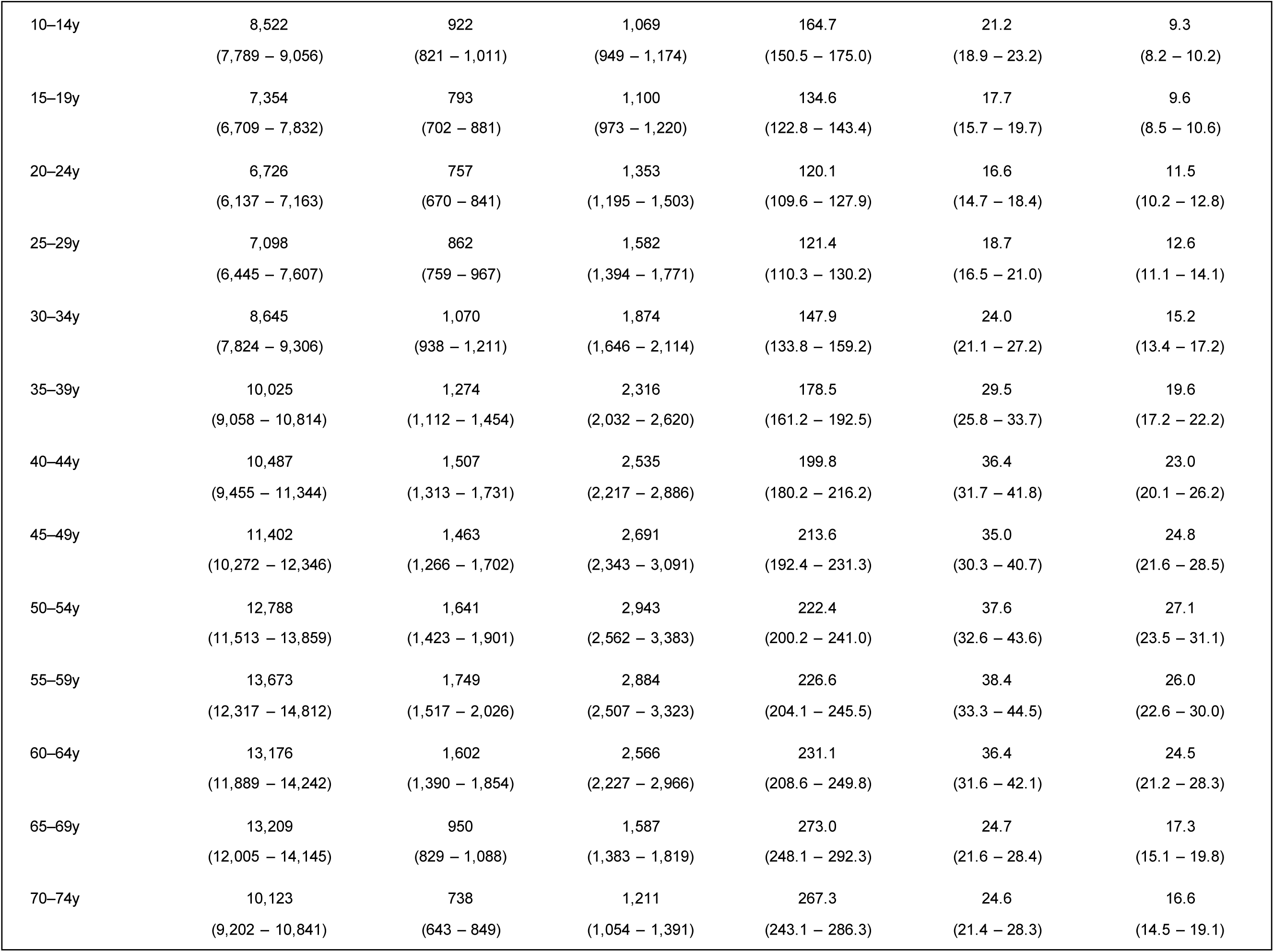
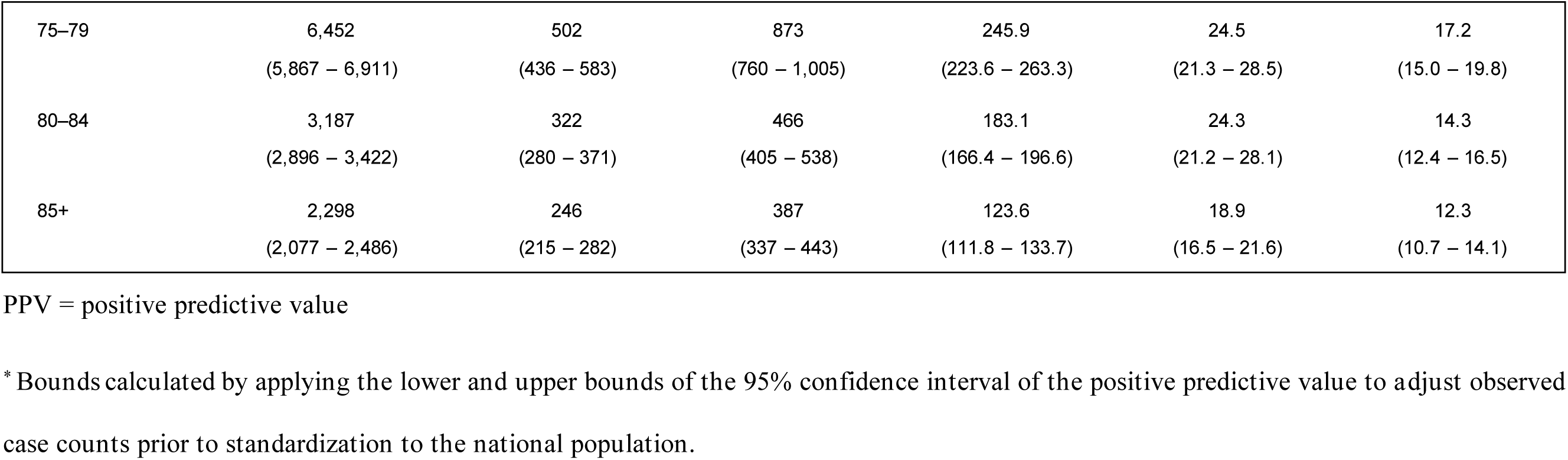
By region: Estimated average annual case count and incidence of Lyme disease per 100,000 population, by stage, sex, and age, 2016-2023.

LD incidence followed a seasonal pattern in all regions, with cases peaking in June and July (Figure 4). This trend was most evident in high-incidence states, with 6.4 times the number of cases in July compared to February. In comparison, neighboring states had 4.9 times more cases and low-incidence states had 2.3 times more cases in July compared to February. Disseminated LD accounted for 22.5% of cases in high -incidence states, 24.6% of cases in neighboring states, and 28.6% of cases in low-incidence states, although, from December through March of each year, over 30% of LD diagnoses were disseminated disease in all regions (Figure 4).

**Figure 4:**
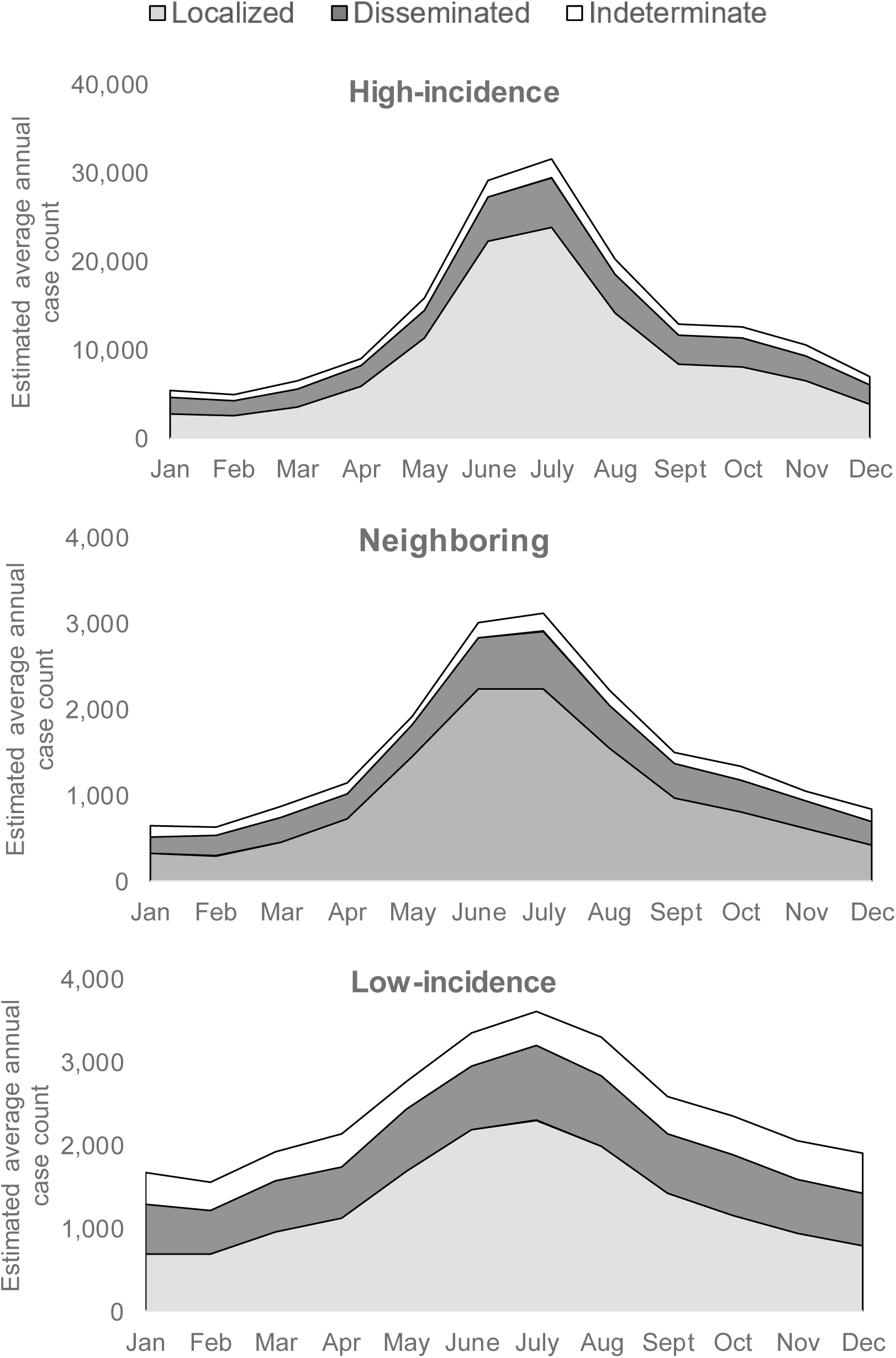
These plots depict the estimated average annual Lyme disease (LD) case count in the US by region, calendar month, and disease stage, from January 2016 through December 2023. Localized, disseminated, and indeterminate disease stage definitions are described in Figure 1. High -incidence LD, neighboring, and low-incidence LD region designations are based on CDC surveillance data from 2021 through 2023.

The average annual incidence of LD ranged from 191.7 new cases per 100,000 population in high -incidence states to 16.6 new cases per 100,000 population in low-incidence states (Table 4). In low-incidence and neighboring states, LD incidence was higher among women than men, while in high -incidence states, incidence was slightly higher among men. This trend was consistent for each year of the study period.

The age distribution of LD incidence varied by region (Table 4; Figure 5). High -incidence states had a clear bimodal distribution of LD incidence by age, with 248 cases per 100,000 individuals among ages 5 – 9 and roughly 270 cases per 100,000 individuals among ages 65 – 74. In neighboring states, LD incidence peaked around 36 cases per 100,000 individuals between ages 40 – 64, with a lower peak for ages 5 – 9. Meanwhile, in low-incidence states, LD incidence peaked at 27 cases per 100,000 individuals between ages 50 – 59 and had only 12 cases per 100,000 individuals for ages 5 – 9. The relative burden of disseminated LD increased with age, particularly in low-incidence and neighboring states, where disseminated disease accounted for over 30% of LD diagnoses for all age categories ≥40 and ≥55 years of age, respectively (Figure 5).

**Figure 5:**
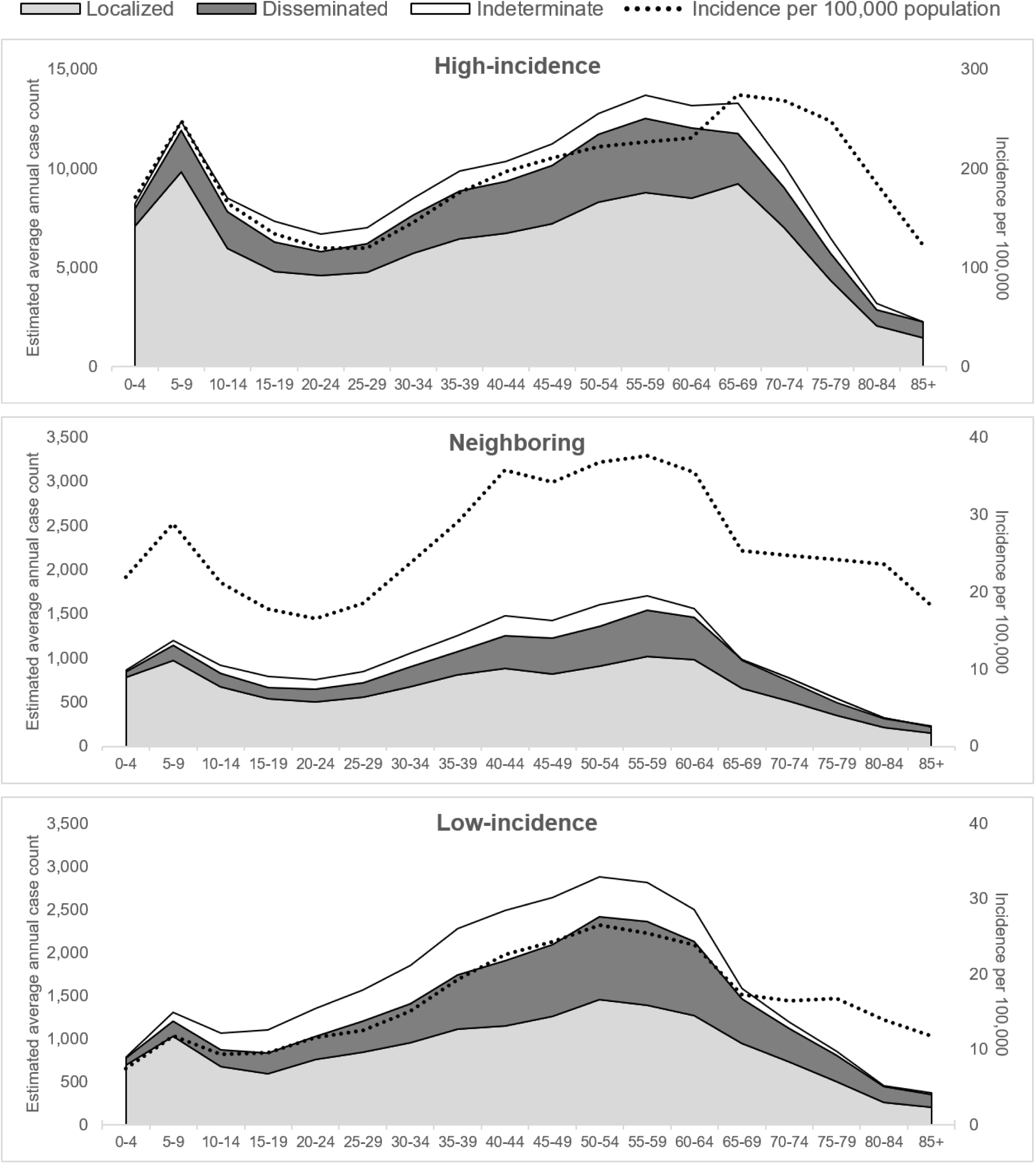
This figure depicts estimated average annual Lyme disease case counts and incidence per 100,000 population in the US, by region, age category, and disease stage. Localized, disseminated, and indeterminate disease stage definitions are described in Figure 1. High-incidence LD, neighboring, and low-incidence LD region designations are based on CDC surveillance data from 2021 through 2023. Note: Y-axis maximum is higher for high-incidence states than neighboring and low-incidence states.

## Discussion

Based on claims data from over 66 million US residents annually, we estimated 212,628 new LD cases each year from 2016 through 2023, among which 66% were classified as localized LD and 24% were classified as disseminated LD. Observed trends by age, sex, and calendar month reflect known patterns.

While prior research has applied algorithms to identify LD in claims data, this is the largest study to date, the first to assess national trends with respect to disease stage at diagnosis, and the first to include populations with both commercial insurance and Medicare. We adjusted claims-based case counts by algorithm- and region-specific PPVs to account for heterogeneity in the performance of these algorithms according to regional disease prevalence. To improve our capture of LD, we added secondary algorithms to identify cases of disseminated LD without an LD diagnosis claim.

The standardized incidence rates we report align with the CDC’s most recent estimates from claims data. Our estimated average annual incidence of 64.6 cases per 100,000 is slightly lower than the estimate by Schwartz and co-authors of 73.3 per 100,000 between 2010 – 2018 (*8*), likely because we adjusted for algorithm PPVs; however, we report comparable trends by sex, age, and region. Our findings also corroborate those of Kugeler and co-authors (*8*), who estimated an average of 205,000 annual LD cases in the US. To adjust for imperfect sensitivity, they multiplied their estimate by a correction factor of 2.33 (= 1/0.43) based on pooled data from three studies of data from 2009 and 2011 that estimated that only 43% of LD cases had an LD diagnosis code, resulting in a final estimate of 476,000 new LD cases in the US annually (*7,24*). We applied a more modest adjustment in our sensitivity analysis based on recent evidence that 61.5% of LD cases may be captured with an LD diagnosis code (*22,23*), resulting in a maximum estimated annual burden of 327,794 cases. Applying the same multiplier as the CDC would have yielded an estimate of 469,712 cases per year, highlighting the importance of evaluating under-diagnosis to accurately measure LD burden.

We expanded on prior studies of US LD incidence by describing the burden of disseminated LD across the US population. Our estimate that 24% of cases are diagnosed during stages of disseminated disease highlights an opportunity for earlier LD detection and treatment, which would reduce the risk of progression to disseminated disease (*25*). We found that low-incidence states had a higher proportion of cases with disseminated disease compared to high-incidence states, although the absolute burden of disseminated disease was higher in high - incidence states. Additionally, the proportion of cases classified as disseminated LD increased with age in all regions. These findings may inform strategies for targeted education related to LD prevention, detection, and treatment for patients and clinicians.

The estimated burden of localized versus disseminated disease from the current study is consistent with a national surveillance report from 2008-2015 that identified a lower burden of erythema migrans (64.7% vs 72.3%) and higher burden of neurologic manifestations (20.0% vs 12.2%) in low-incidence compared to high-incidence states (*10*). However, the national surveillance report lacked clinical data for nearly 40% of reported cases, and did not collect data on non-specific localized symptoms, as national surveillance is not intended for disease staging. Current national surveillance following the 2022 LD case definitions captures no clinical data in high-incidence states as it relies solely on laboratory data in these states (*26*). These limitations of surveillance data reinforce the need for an alternate data source to characterize the national burden of LD.

This study had several limitations. With no direct data from claims on the sensitivity of the primary LD algorithm, we used two strategies to adjust for sensitivity - identifying additional cases based on secondary algorithms, and applying a correction factor - both of which have limitations. Supplementing LD capture with secondary algorithms for disseminated LD cases without a LD diagnosis code fails to capture the population of localized LD cases without a diagnosis code. Meanwhile, the correction factor, designed to account for both localized and disseminated disease without a LD diagnosis, was calculated from two studies using electronic health records. Sensitivity of the primary LD algorithm in electronic health records may differ from sensitivity of the algorithm in claims data. Additionally, while the primary and secondary algorithms we used to identify LD cases have been validated in high-incidence and neighboring states, they have not been validated in low-incidence states. We applied the PPVs from neighboring states (i.e., those bordering high incidence states), and in a sensitivity analysis, also applied lower PPVs (50% and 20%), to low-incidence states. The results of these sensitivity analyses can provide reasonable bounds on the true total burden of LD in low-incidence states but further work is needed to more accurately quantify disease incidence in these regions. Uncertainty regarding the total burden of disease should not diminish the trends we observed in low-incidence states by sex, age, stage, and over time. Additionally, we recognize that the assumption of 100% PPV for the unvalidated inpatient algorithm is not accurate, although inpatient codes typically have higher PPVs than outpatient codes (*14–16*). Knowing that this algorithm also has imperfect sensitivity, and that inpatient LD diagnoses accounted for only 1.7% of adjusted cases annually, we do not consider this assumption to be a major source of error in our analysis. The staging criteria we applied to categorize cases meeting the primary algorithm as localized, disseminated, or indeterminate have also not been validated, although they have been used previously (*17,18*). Finally, while we included a very large national sample, we did not have data for individuals on Medicaid or those who are uninsured. If LD incidence in these populations differs systematically compared to the population we included, then our estimated national incidence rates and national case counts may be under- or over-estimates.

## Conclusion

Our results corroborated prior estimates of the national burden of Lyme disease in the US and shed light on patterns in the incidence of localized and disseminated LD. The substantial burden of disseminated LD that we observed underscores the need for improved detection and treatment of early LD.

## List of abbreviations

CDC: Centers for Disease Control and Prevention
CI: Confidence interval
CMS: Centers for Medicare & Medicaid Services
IV: Intravenous
LD: Lyme disease
PPV: Positive predictive value
US: United States

## Declarations

### Ethics approval and consent to participate

The Harvard Pilgrim Health Care IRB determined this project did not meet the definition of human subject research under the purview of the IRB and consent to participate was deemed unnecessary according to federal HHS regulations 45 CFR 46 (IRB reference #2270365). This study adhered to the Declaration of Helsinki.

### Consent for publication

Not applicable

### Availability of data and materials

The datasets generated and analyzed during the current study are not publicly available due to data-sharing restrictions among the insurers that contributed data for this study.

### Authors’ contributions

Study concept and design: SAK, SJW, JHS, NMC. Acquisition of data: JO, KS, KC, DE, ER, RJ, KD, SML, DAD, MS, AD, QM. Analysis and interpretation of the data: SAK, SJW, JA, JHS, JCM, NMC. Drafting of the manuscript: SAK, SJW, JHS, NMC. Critical review of the manuscript: All authors.

## Acknowledgements

The authors would like to thank Justin O’Hagan and Rebecca Hawrusik for their critical review of the manuscript.

## Appendices

**Appendix Table 1.**
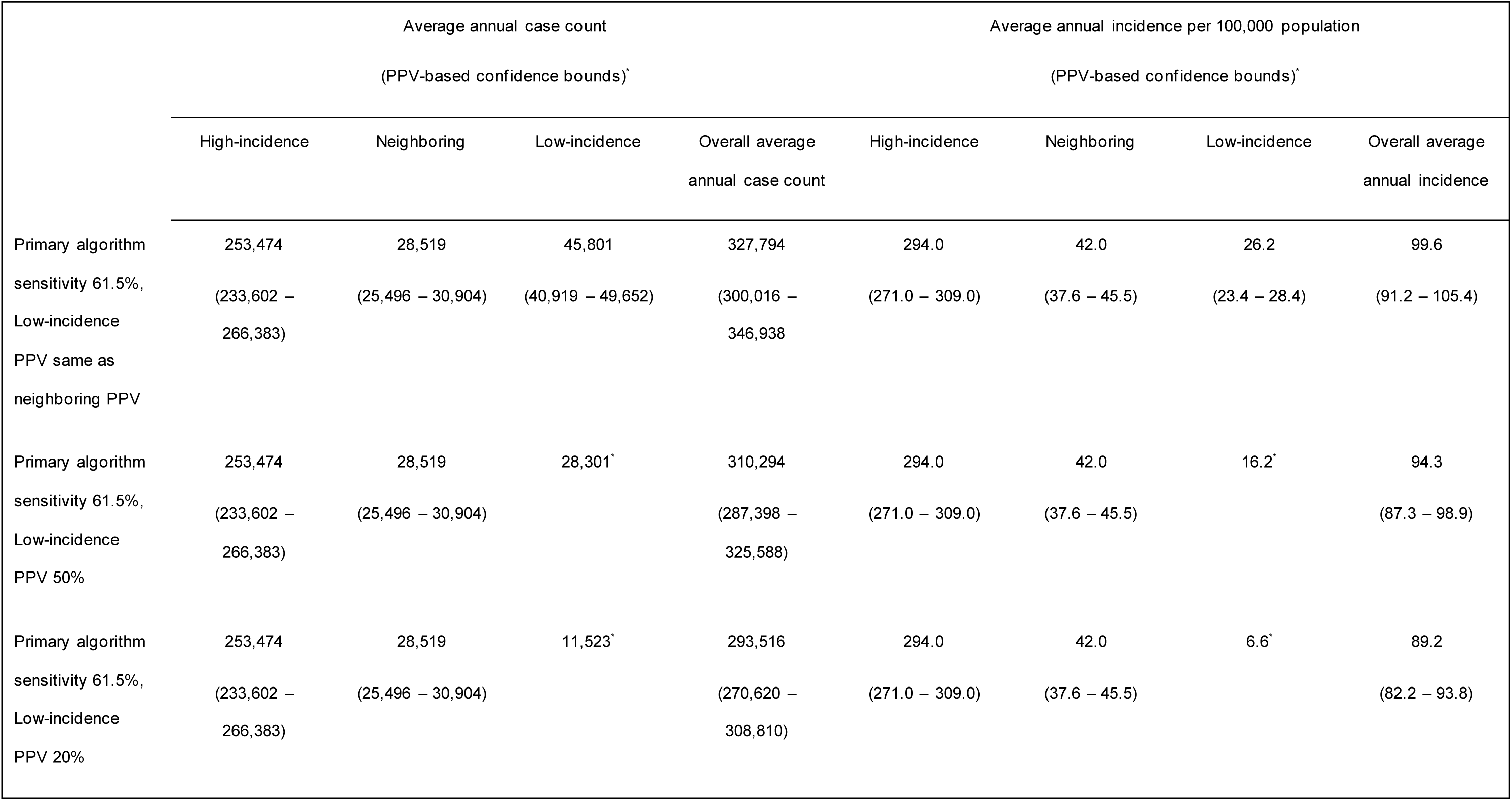
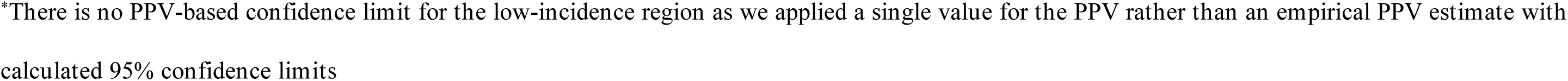
Sensitivity calculations of average annual case count and incidence of Lyme disease per 100,000 population overall and by region, 2016 – 2023.

